# A novel cuproptosis-related gene signature for the prediction of liver cancer prognosis identified DLAT is a potential therapeutic target

**DOI:** 10.1101/2022.10.23.22280648

**Authors:** Dong Xu, Fei Yang, Yang Chen, Hong Zhu, Haijian Sun, Tong Shen, Yongkang Zhu, Guowei Zhou, Dexuan Chen, Xiaojun Yang, Kai Lin, Cunbing Xia

## Abstract

**Background and Purpose:** The liver cancer (LC) is a highly malignant tumor of the digestive system with a poor prognosis. Cuproptosis is a new type of regulated cell death that has been found by researchers. The expression of cuproptosis-related genes in LC and their relevance to prognosis, on the other hand, remain unknown. This study aimed to explore a gene signature to predict the liver cancer prognosis and identified the vital gene.

**Experimental approach:** The expression patterns of RNA and related clinical data of 371 LC patients were obtained based on The Cancer Genome Atlas (TCGA). Differentially expressed genes (DEGs) were acquired by comparing tumors with adjacent normal samples. Genes displaying significant association with OS were screened through univariate Cox regression analysis and the least absolute shrinkage and selection operator (LASSO) algorithm. All cases were classified into the validation or training group to validate the constructed gene signature. We conducted real-time polymerase chain reaction (PCR) and assays for transwell invasion, CCK-8, and colony formation to determine the biological roles of DLAT.

**Key Results:** The differential expression of twelve cuproptosis regulators in LC and normal liver tissues was discovered in this investigation. DEGs can be used to distinguish between two forms of LC. Cuproptosis-related genes were evaluated for survival predictive significance using the Cancer Genome Atlas (TCGA) cohort. A 3-gene signature based on least absolute shrinkage and selection operator (LASSO) Cox regression was used to categorize an LC patient cohort from the TCGA into low- and high-risk categories. Patients in the low-risk group had a considerably higher likelihood of surviving (P = 0.05) than those in the high-risk group. When paired with clinical parameters, risk score was an independent predictor in predicting the OS of patients with LC.

**Conclusions & Implications:** Cuproptosis-related genes thus play an important role in tumor formation and can be used to predict the prognosis of LC patients. DLAT has the best prognostic value and can be a therapeutic target for liver cancer.

## Introduction

Liver cancer (LC) is still a global health issue, with an increasing incidence and the fourth biggest cause of cancer-related death worldwide[1]. LC has the highest incidence and mortality rates in East Asia and Africa, while it is also increasing in certain parts of Europe and the United States[2]. Indeed, according to Surveillance Epidemiology End Results (SEER), LC has been the fastest-growing cause of cancer-related death in the United States since the early 2000s, and if current trends continue, LC will become the third greatest cause of cancer-related death by 2030.[3]. Because LC is a distinct neoplasm that develops in individuals with cirrhosis in 80–90% of instances, the utilization of various treatment alternatives may be limited according to the patient’s general health situation. Patients with early-stage tumors are the best prospects for excision, transplantation, and local ablation, while patients with intermediate-stage tumors are the best candidates for TACE, and those with advanced disease will undergo systemic therapy first. Despite recent advances in therapy, the 5-year survival rate has remained stagnant. Given the limits of current LC treatments, new therapeutic targets are necessary to enhance the clinical outcome of the disease; hence, accurate new prognostic models are required immediately to make targeted therapies more practical. Tsvetkov and colleagues have found a new type of controlled cell death called cuproptosis, which is distinct from oxidative stress-related cell death (e.g., apoptosis, ferroptosis, and necroptosis). Excessive intracellular copper concentrations cause cell death, according to Tsvetkov et al.[4]. They discovered that cells that rely heavily on mitochondrial respiration are more susceptible to copper-induced cell death, implying a link to the TCA cycle. They discovered the key genes involved in copper-induced cell death, including FDX1, as well as six other genes that encode lipoic acid pathway-related enzymes, such as lipolytransferase 1 (LIPT1), lipoyl synthase (LIAS), and dihydrolipoamide dehydrogenase (DLD), as well as protein targets of lipoylation, such as pyruvate dehydrogenase (PDH) complex, including FDX1 has the ability to control DLAT lipoylation. Furthermore, copper can stimulate DLAT oligomerization, resulting in an increase in insoluble DLAT, which leads to proteotoxic stress and cell death. More and more evidences indicate that copper ion and copper ion related genes play an important role in the development of LC. The content of copper ion is closely related to the risk of liver cancer. Copper ions can mediate the occurrence of hepatitis and liver cirrhosis, and then cause the occurrence of liver cancer through reactive oxygen species (ROS) [5–8]. And it is reported that one of the cuproptosis-related gene FDX1 is clearly associated with the risk of hepatocellular carcinoma [8]. Therefore, cuproptosis-related genes have the potential to predict the occurrence and development of liver cancer, which is expected to be a target for liver cancer treatment in the future.

Cuproptosis has a crucial part in the formation of tumors and antitumor activities, according to available evidence; nevertheless, its precise roles in LC have not been investigated. As a result, we conducted a thorough investigation to compare the expression levels of cuproptosis-related genes in normal liver and LC tissues, investigate their predictive significance, and investigate the relationships between cuproptosis and the tumor immune microenvironment.

## Results

### Identification of DEGs between LC tumor and normal tissues

The expression levels of 13 cuproptosis-related genes were compared in data from 50 normal and 374 tumor tissues from The Cancer Genome Atlas (TCGA), and 12 differentially expressed genes (DEGs) were discovered (all P < 0.05). Four genes were downregulated (DBT, SLC31A1, GCSH, and FDX1), while eight others were enriched in the tumor group (DLST, DLD, PDHB, PDHA1, LIAS, LIPT1, DLAT, and ATP7A). Figure 1A shows heatmaps depicting the RNA levels of these genes (green: low expression level; red: high expression level). The results of a protein–protein interaction (PPI) analysis was used to further investigate the interactions of these cuproptosis-related genes, as shown in Fig. 1B. We identified that LIAS, DLST, GCSH, DLAT, LIPT1, DLD, PDHB, PDHA1, DBT, SLC31A1, ATP7A, and ATP7B were hub genes by setting the minimum required interaction score for the PPI analysis to 0.9 (the highest confidence). Figure 1C depicts a correlation network that includes all cuproptosis-related genes (red: positive correlations; blue: negative correlations).

**Figure 1.**
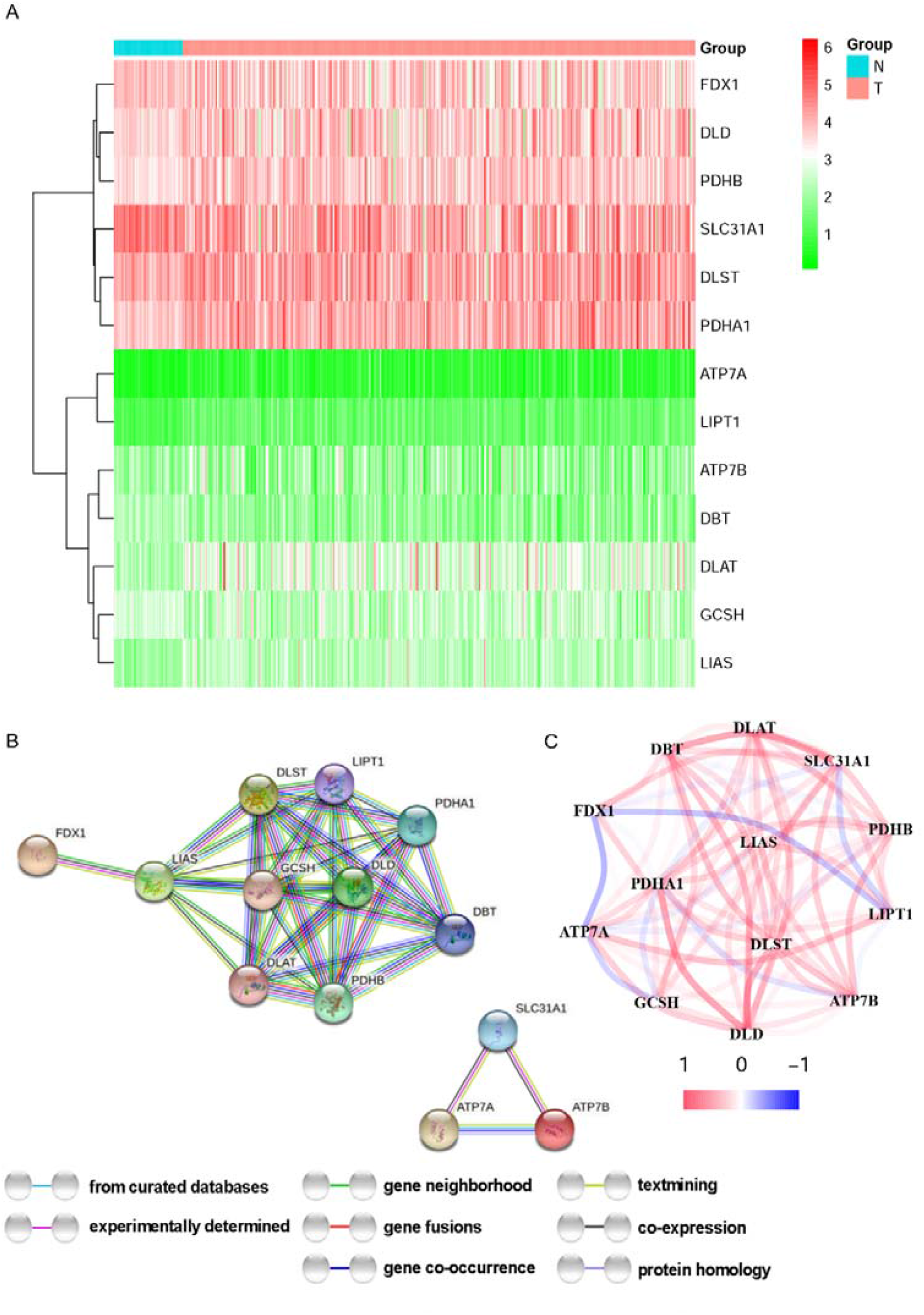
Expressions and interactions of the 13 cuproptosis-related genes. **A** Heatmap (green: low expression level; red: high expression level) of the cuproptosis-related genes in normal (N, brilliant blue) and tumor tissues (T, red). **P < 0.01; ***P < 0.001 were used to represent P values. **B** PPI network displaying cuproptosis-related gene connections (interaction score = 0.9). **C** The cuproptosis-related genes’ association network (red line: positive correlation; blue line: negative correlation). The strength of the relevance is shown in the color depth.

### The TCGA cohort was used to develop a prognostic gene model

A total of 374 LC samples were matched to patients with complete survival data. The survival-related genes were first screened using univariate Cox regression analysis. The three genes (DLAT, LIPT1, and ATP7A) that met the P < 0.01 threshold were kept for additional investigation, and all of them were linked to an increased risk with HRs >1 (Fig. 2A). The ROC curve of each gene was shown in Figure S2. A 3-gene signature was generated based on the LASSO Cox regression analysis (Fig. 2B, C). The area under the ROC curve (AUC) of ATP7A was 0.66 for 1-year, 0.561 for 2-year and 0.563 for 3-year. The AUC of DLAT was 0.695 for 1-year, 0.627 for 2-year and 0.609 for 3-year. The AUC of LIPT1 was 0.638 for 1-year, 0.558 for 2-year and 0.588 for 3-year. Collectively, DLAT has the best prognostic value for 1, 2 and 3 years. Risk score = (0.3005*LIPT1 exp.) + (0.06026*DLAT exp.) + (0.21313*ATP7A exp.) 374 patients were stratified into low- and high-risk categories based on the median score determined by the risk score method (Fig. 2D). Patients with varied risks were well segregated into two clusters using principal component analysis (PCA) (Fig. 2E). Patients in the high-risk group died more frequently and lived for a shorter time than patients in the low-risk group (Fig. 3F, on the right side of the dotted line). The low-risk and high-risk groups had a significant difference in OS time (P < 0.05, Fig. 2G). The area under the ROC curve (AUC) was 0.722 for 1-year, 0.609 for 2-year, and 0.620 for 3-year survival, according to time dependent receiver operating characteristic (ROC) analysis (Fig. 2H). We also validated the prognostic value of the signature with 70% randomly drawn from the TCGA cohort. The result was consistent with the TCGA cohort (Fig. S1). The time-dependent ROC curve showed the AUCs was 0.769 at 0.778 for 1-year, 0.626 for 2-year and 0.602 for 3-year.

**Figure 2.**
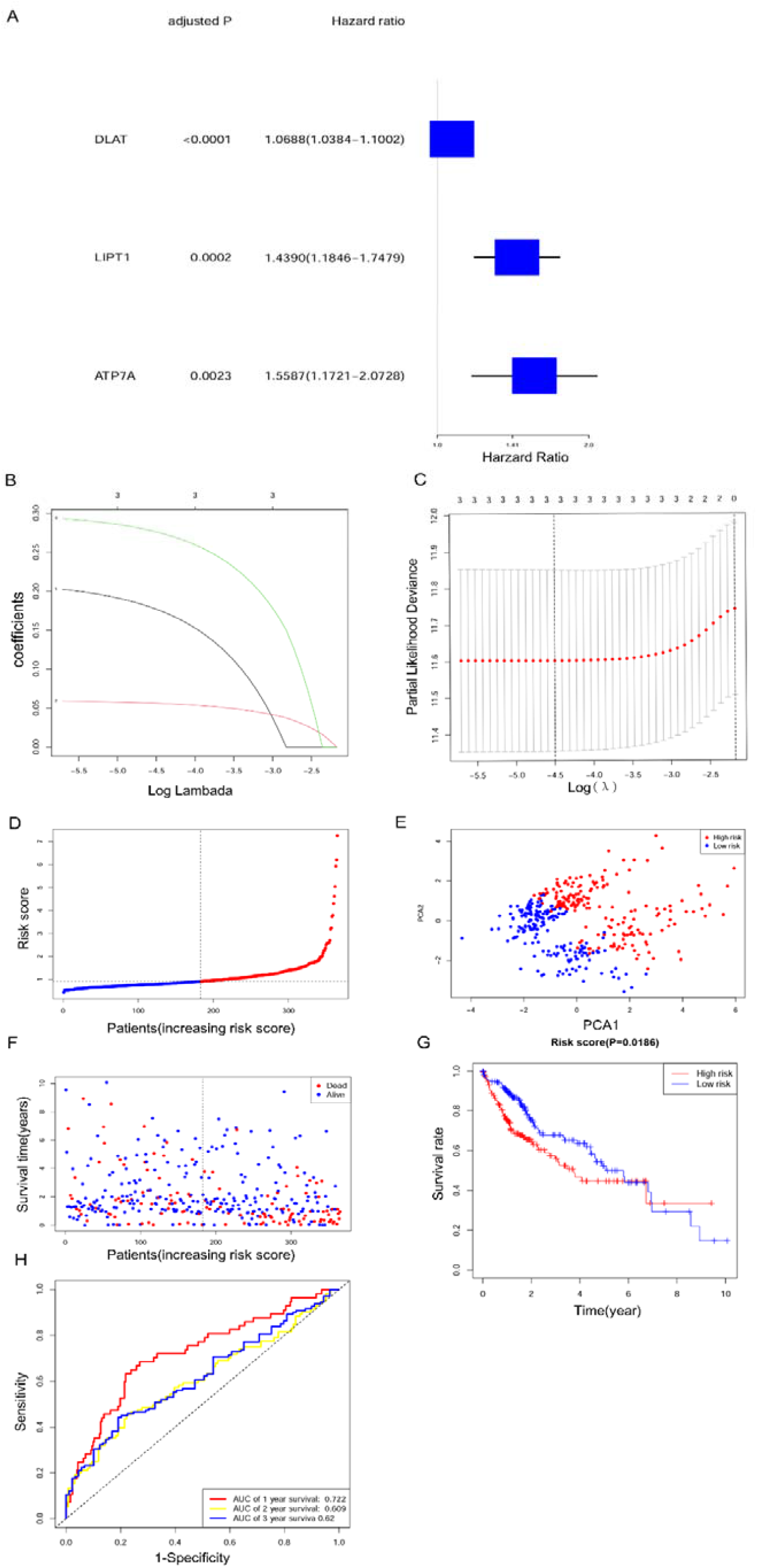
TCGA cohort risk signature construction. **A** For each cuproptosis-related gene, a Univariate cox regression analysis of OS was performed, and three genes with P < 0.01 were found. **B** LASSO regression of the three genes linked to OS. **C** In the LASSO regression, cross-validation is used to fine-tune the parameter selection. **D** Patients are assigned to groups depending on their risk scores. Based on the risk score, **E** PCA plot for LCs. **F** Each patient’s chance of survival (low-risk population: on the left side of the dotted line; high-risk population: on the right side of the dotted line). **G** Kaplan–Meier curves for the OS of high-risk and low-risk individuals. **H** The risk score’s prediction efficiency was proved by ROC curves.

**Figure 3.**
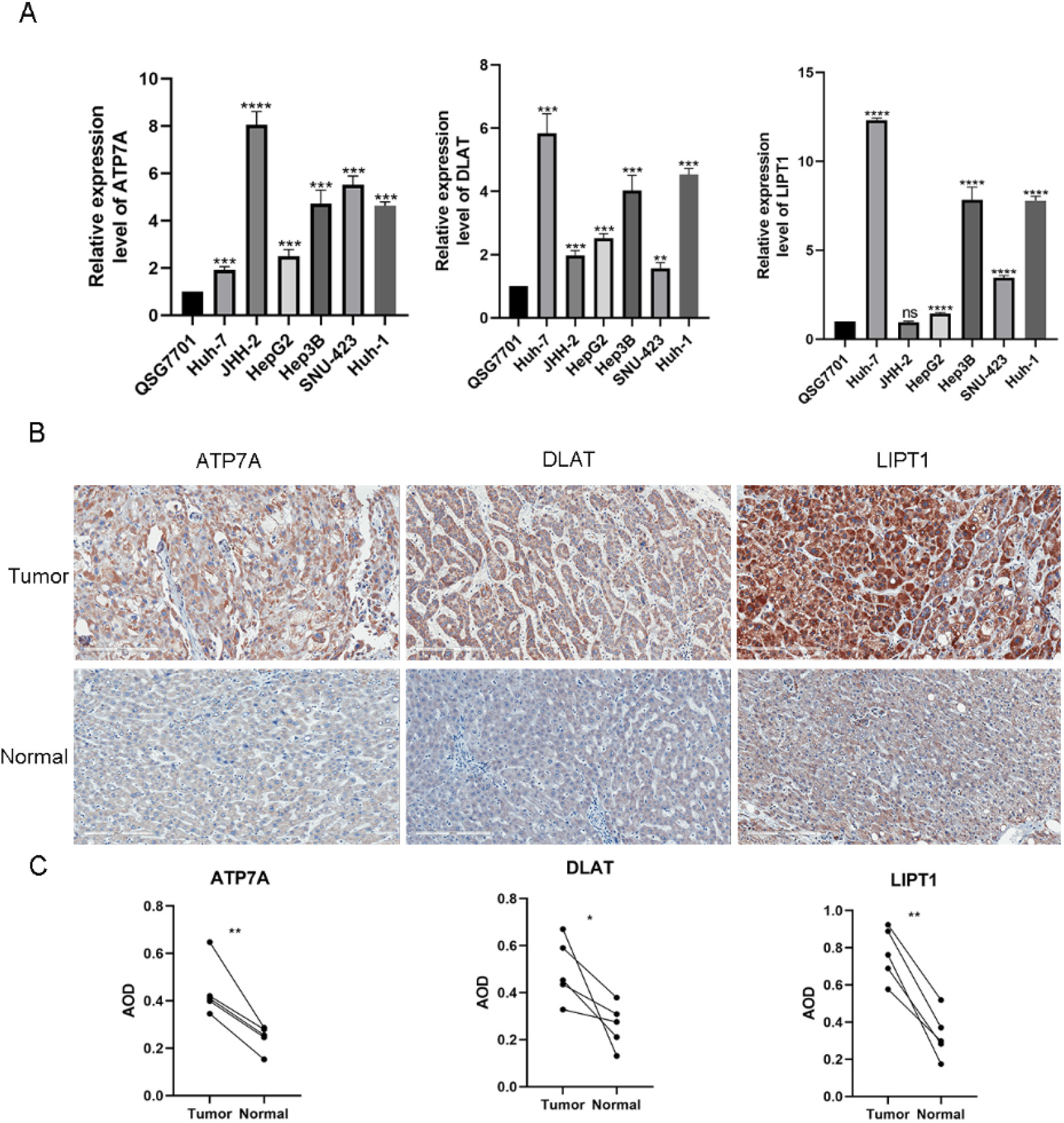
Expression of and alterations in the three DEGs. **A** Relative mRNA expression level of ATP7A, DLAT and LIPT1 in JHH-2, SNU-423, Huh-1, HepG2, Huh-7, Hep3B and QSG7701 by PCR (2^−ΔΔct^). **B** The protein expression analysis by IHC.

### Validation of the expression of and alterations in the three genes

The mRNA expression levels (2^−ΔΔct^) of the three genes (ATP7A, DLAT and LIPT1) were validated using PCR in liver cancer cell lines (JHH-2, SNU-423, Huh-1, HepG2, Huh-7 and Hep3B) and normal liver cell line (QSG7701) (Fig. 3A). Liver cancer cell lines show a higher expression level of three genes than normal liver cell line. And IHC showed the same result in patients’ tissues (Fig. 3B, C).

### The risk model’s independent prognostic value

To see if the risk score produced from the gene signature model could be utilized as an independent prognostic factor, we used univariate and multivariable Cox regression analysis. The risk score was found to be an independent predictor of poor survival in the TCGA cohorts in a univariate Cox regression analysis (HR = 1.5142, 95% CI: 1.0581–2.1670, Fig. 4A). After adjustment for other confounding factors, the multivariate analysis revealed that the risk score was a prognostic predictor (HR = 1.4936, 95% CI: 1.0418– 2.1411, Fig. 4B) for patients with LC. In addition, for the TCGA cohort, we created a heatmap of clinical characteristics (Fig. 4C).

**Figure 4.**
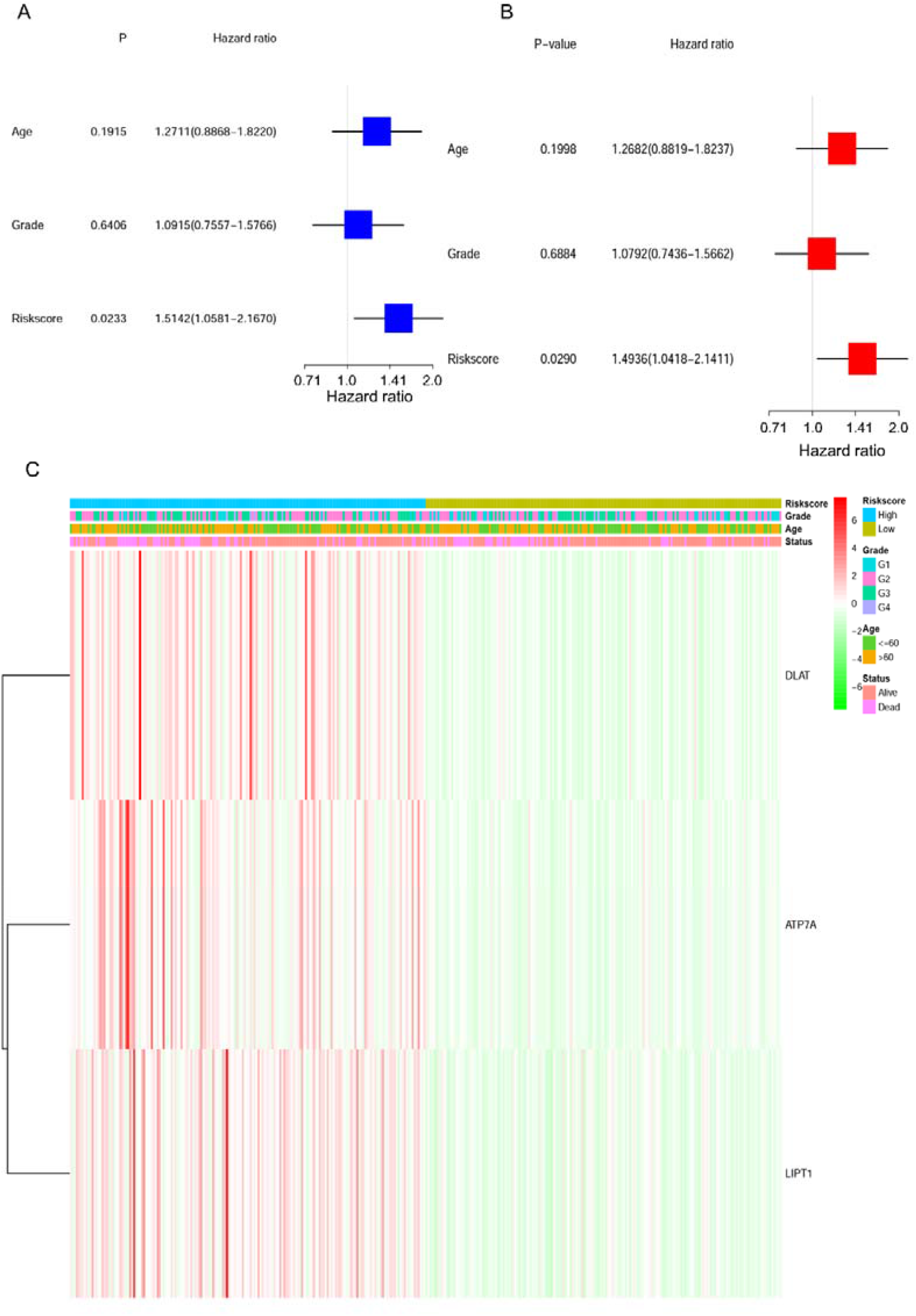
Univariate and multivariate Cox regression analyses for the risk score. **A** For the TCGA cohort, a univariate analysis (grade: the degree of tumour differentiation, G1 to G4). **B** The TCGA cohort’s multivariate analysis **C** Connections between clinicopathologic characteristics and risk groups (P < 0.05) heatmap (green: low expression; red: high expression).

### Association between the signature and clinical characteristics

Chi-square test was used to explore whether the prognostic signature participated in the development and progression of LC. The result (Fig. 5) showed that there were significant differences between high- and low-risk groups in tumor grade (p=0.012), TNM stage (p=0.003), and T stage (p=0.006). Moreover, stratification analysis was further conducted to investigated the prognostic significance of the signature in subgroups. Our research suggested that DEG-based signature showed excellent performance in predicting outcome in male (p=0.007) and high-risk group (p=0.047). While DEG-based signature showed poor performance in predicting outcome in other groups (p > 0.05) (Fig. 6).

**Figure 5.**
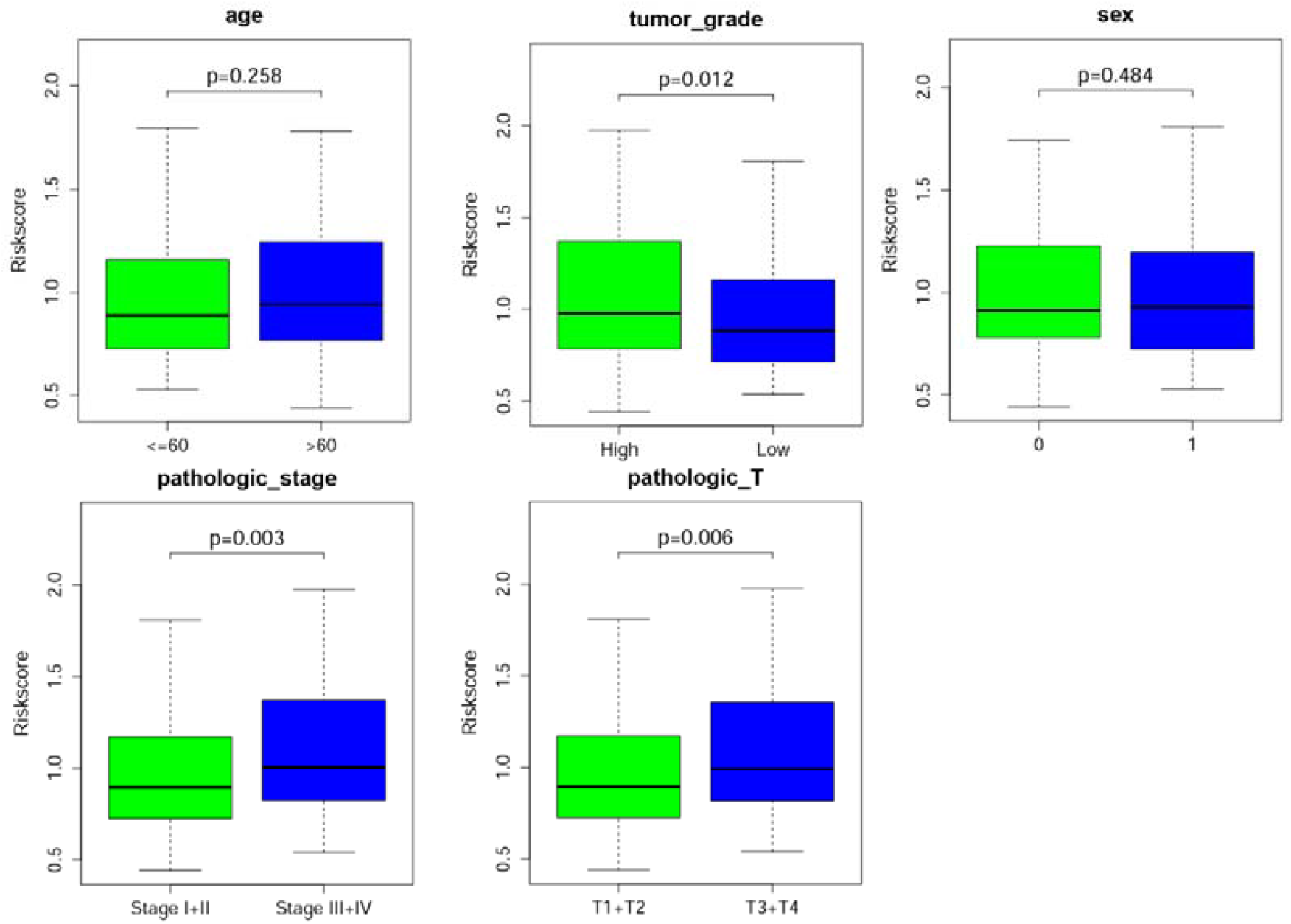
Correlation between signature and clinical characteristics.

**Figure 6.**
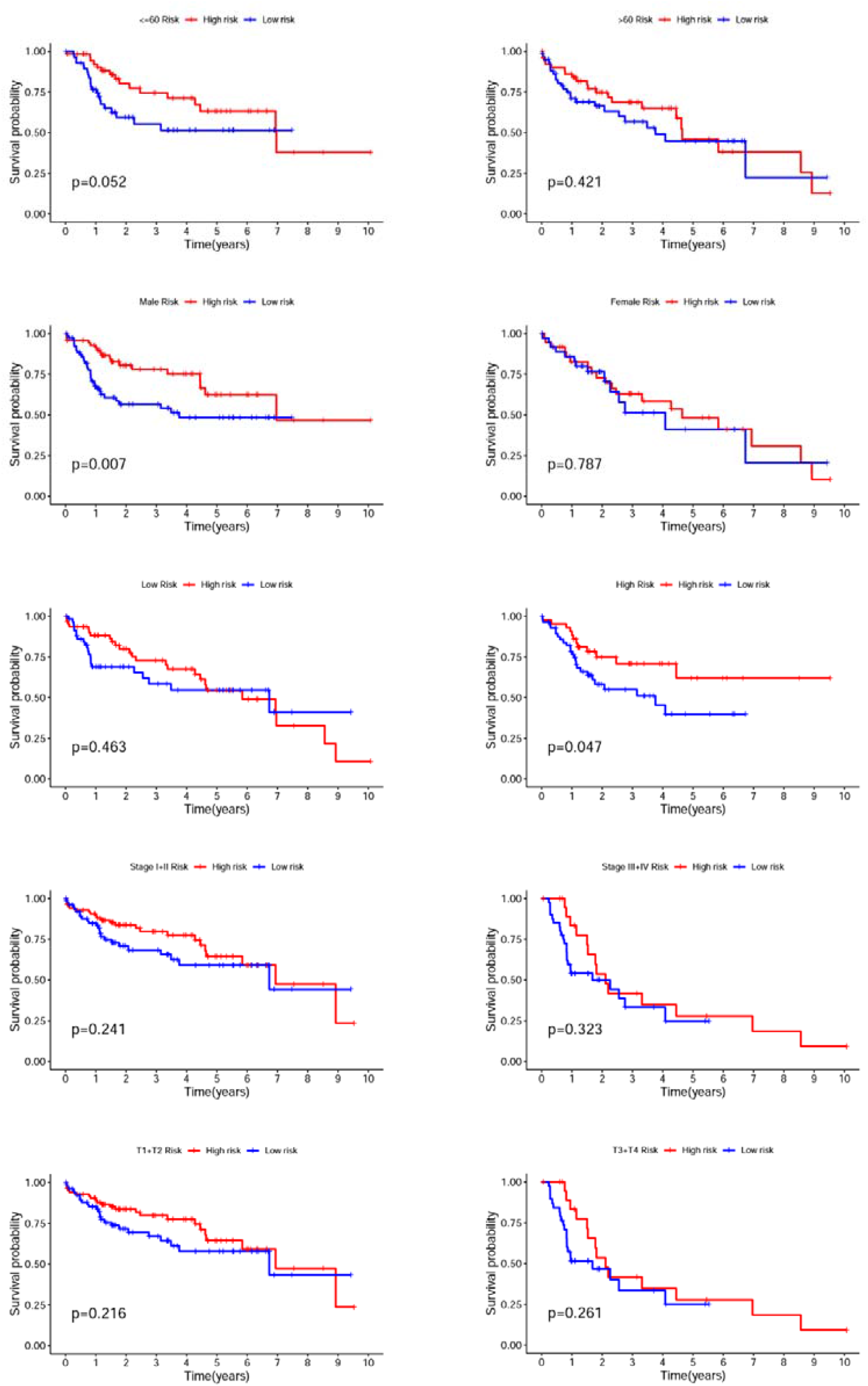
Kaplan-Meier curves of OS diferences stratifed by age, sex, risk group, tumor stage and T stage between the high-risk groups and low-risk groups.

### Construction of a Nomogram

The nomogram incorporated various prognostic indicators to assess the survival probability of an individual graphically. To further forecast the survival of LC patients, we structured a nomogram comprised of sex, tumor grade, tumor stage and risk score. Nomography predicted the 1-, 3-, 5-year survival rate of patients with LC (Fig. 7A). The calibration curve indicated that the practical survival of the patient was in line with the predicted value (Fig. 7B). The C index of the nomogram was 0.636, which confirmed the favorable prediction ability of the nomogram.

**Figure 7.**
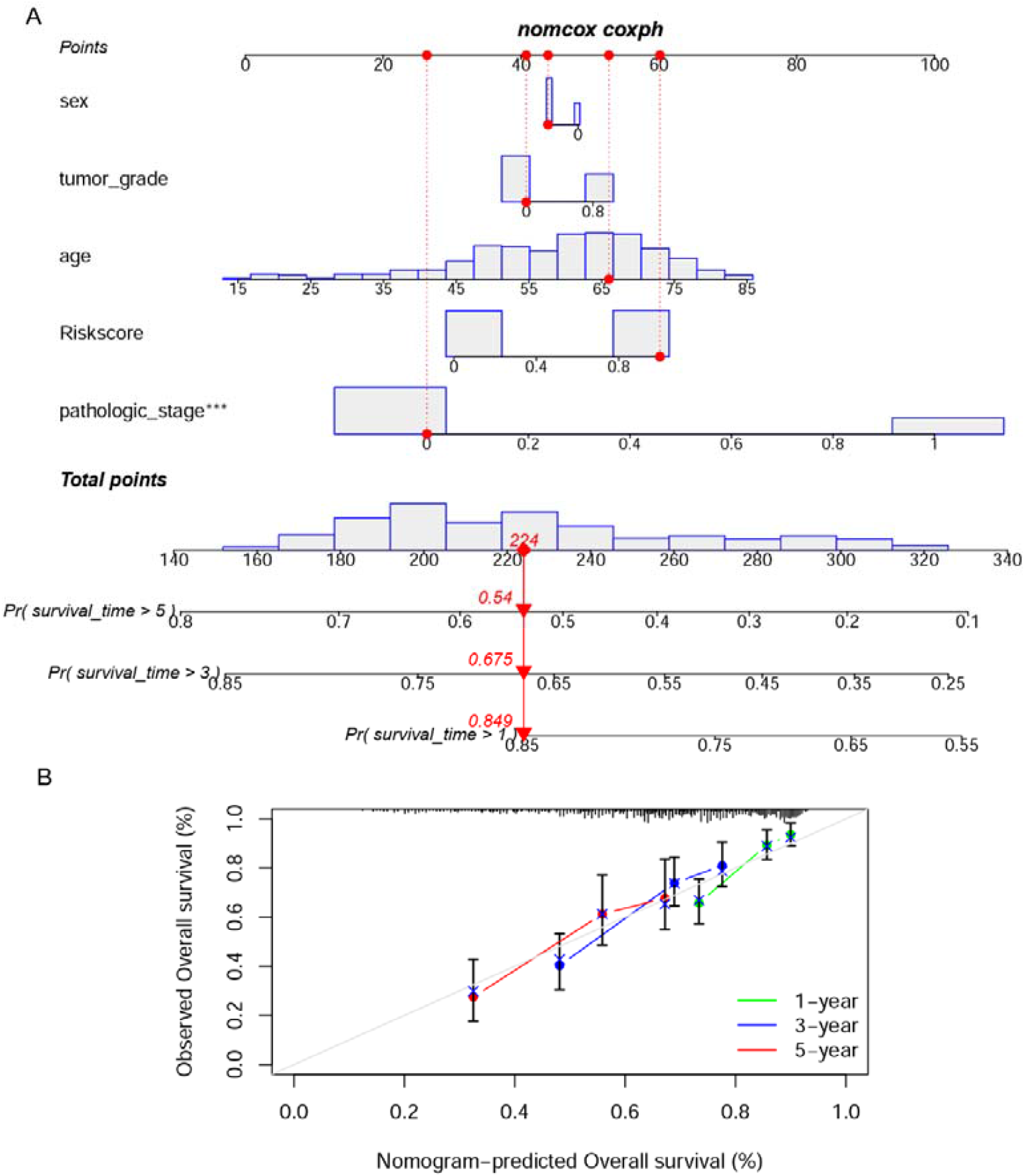
Construction of a nomogram. **A** nomogram for predicting 3- or 5-year OS. **B** The calibration plots for predicting 1-, 3-, 5-year OS.

### Using the risk model to conduct functional assessments

We used the “limma” R package to extract DEGs and applied the FDR < 0.05 and |log_2_FC| ≥ 1 criteria to further investigate the variations in gene functions and pathways between the risk model subgroups. In the TCGA cohort, 365 DEGs were discovered between the low- and high-risk categories. In the high-risk group, 183 genes were elevated, while the other 182 genes were downregulated. These DEGs were then used to undertake gene ontology (GO) enrichment analysis and Kyoto Encyclopaedia of Genes and Genomes (KEGG) pathway analysis. The DEGs were mostly connected with extracellular matrix architecture, channel function, and tumor-associated pathways, according to the findings (Fig. 8A, B).

**Figure 8.**
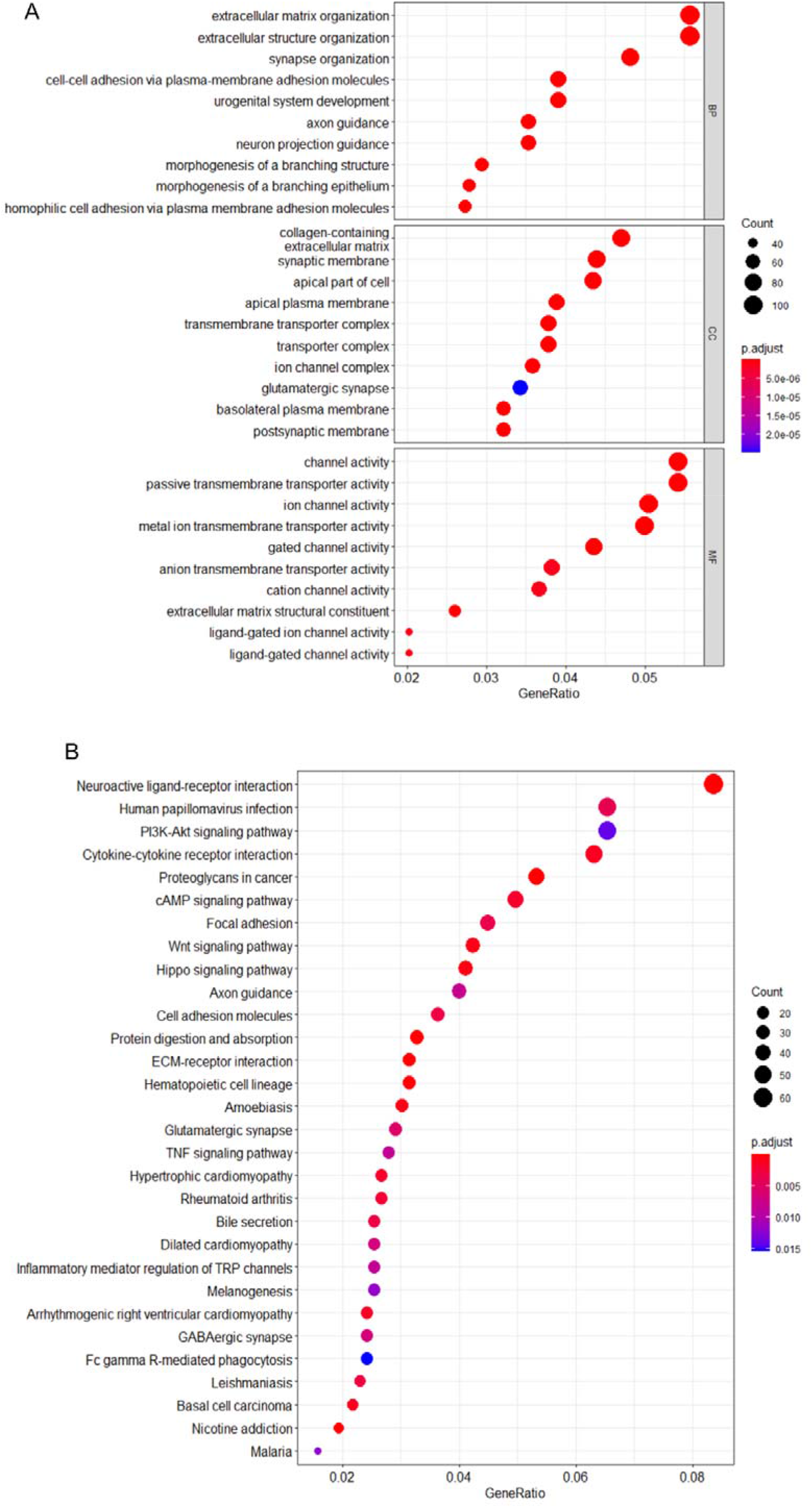
Functional analysis of the DEGs in the TCGA cohort comparing the two risk categories. **A, B** GO and KEGG enrichment bubble graph (the bigger bubble means the more genes enriched, and the increasing depth of red means the differences were more obvious; q-value: the adjusted p-value).

### Comparing immunological activation in different subgroups

Using single-sample gene set enrichment analysis, we examined the enrichment scores of 16 types of immune cells and the activity of 13 immune-related pathways between the low and high-risk groups in the TCGA cohorts based on the functional analyses (ssGSEA). Mast cells and NK cells were found to be lower in the high-risk subgroup than in the low-risk category (Fig. 9A). APC co-stimulation and MHC class I pathway activity were higher in the high-risk group than in the low-risk group in the TCGA cohort, except for cytolytic activity (Fig. 9B).

**Figure 9.**
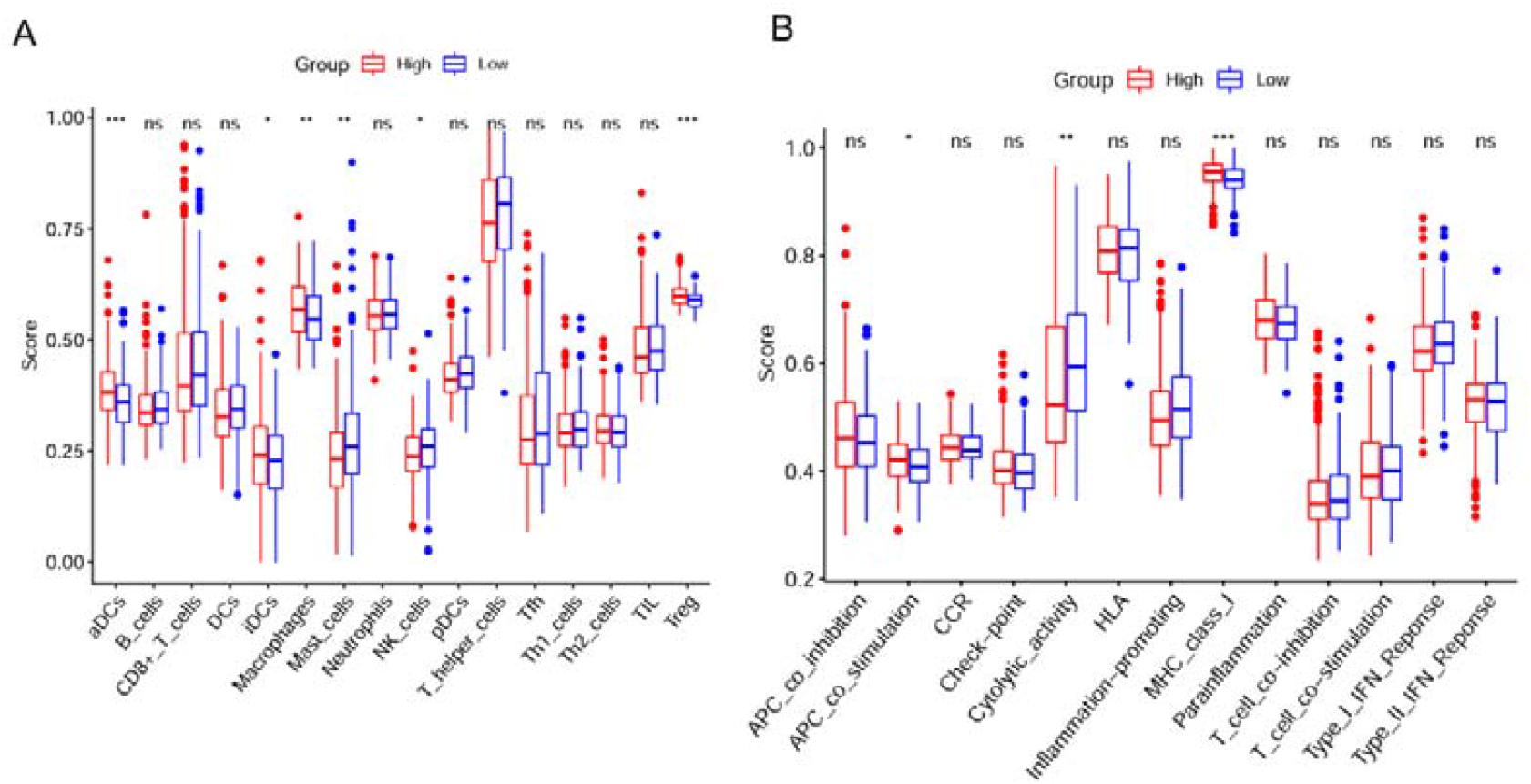
ssGSEA scores for immune cells and immunological pathways compared. **A, B** In the TCGA cohort, the enrichment scores of 16 types of immune cells and 13 immune-related pathways were compared between the low-risk (blue box) and high-risk (red box) groups.

### Immune infiltration level analysis of the DEG-based signature

The relationship between the signature and immune infiltration was displayed in the heatmap according to the analyses of TIMER, CIBERSORT, CIBERSORTABS, XCELL, QUANTISEQ, EPIC, and MCP-counter (Fig. S3). Given the significance of checkpoint inhibitor immunotherapies, we also investigated the correlation between risk score and key immune checkpoints (Additional file 3). We found a prominent difference in the expression of VTCN1, TNFSF18, TNFSF15, TNFSF9, TNFSF4, TNFRSF9, TNFRSF4, NRP1, LGALS9, LAIR1, ICOS, HHLA2, HACVR2, CTLA4, CD276, CD274, CD200R1, CD200, CD86, CD80 and CD44 between the two groups of patients (Fig. 10 and Fig. S4).

**Figure 10.**
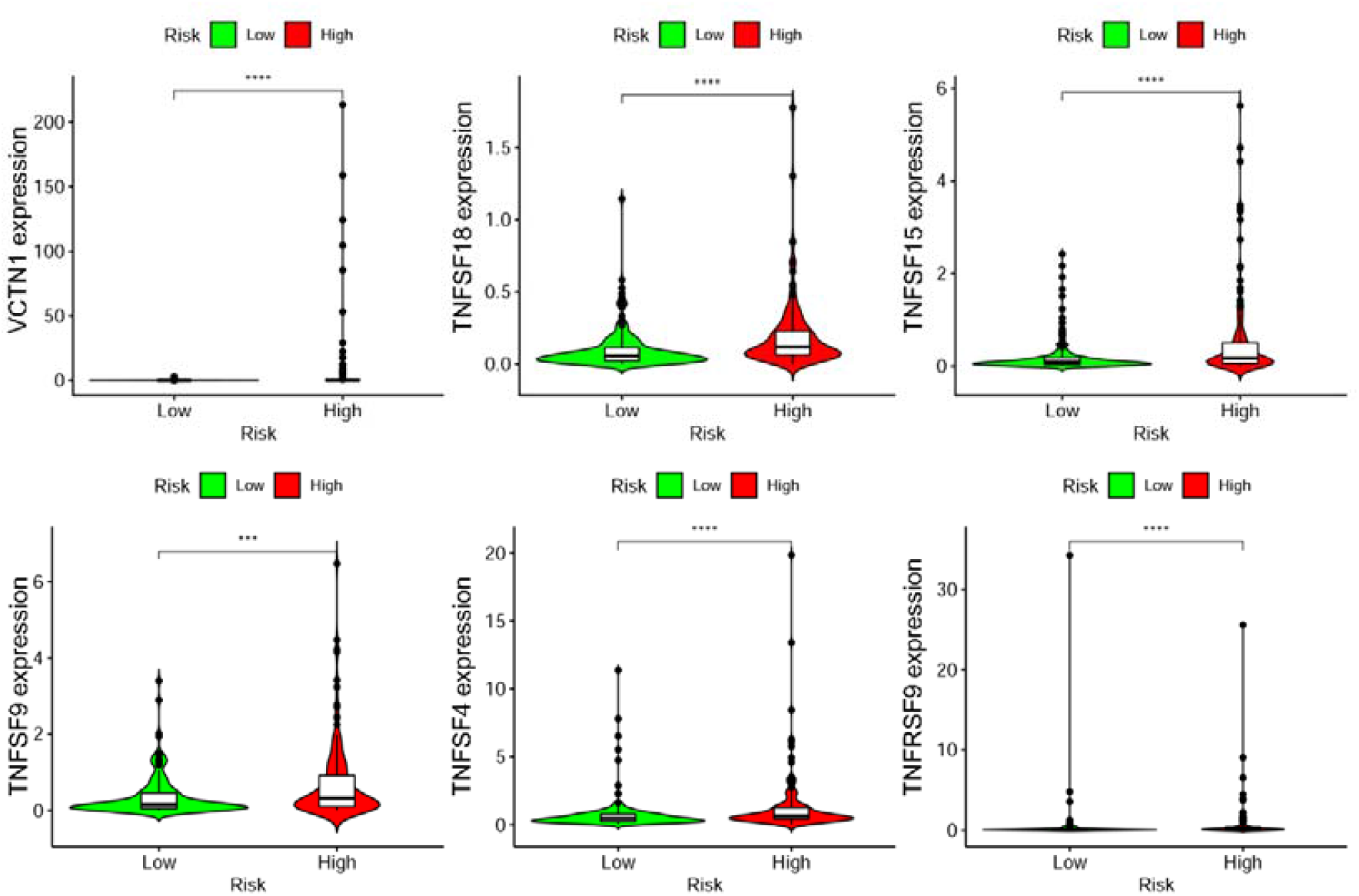
The relationship between prognostic signature and immune checkpoints.

### Identification of small molecule drugs

Enrichr database was used to identify the potential small molecular drugs on the basis of DEGs. The top 5 were vitinoin, latamoxef, chlorzoxazone, atrazine and medrysone (all p < 0.05, Table 1).

**Table 1.** The 5 small molecular drugs of Enrichr dataset analyses results

| Index | Name | P-value | Ajusted<br>p-value | Odds<br>Ratio | Combined<br>score |
| --- | --- | --- | --- | --- | --- |
| 1 | Vitinoin CTD 00007069 | 0.00000009 | 0.000028 | 34.75 | 563.85 |
| 2 | latamoxef HL60<br>DOWN | 0.00001044 | 0.001602 | 16.41 | 188.25 |
| 3 | chlorzoxazone HL60<br>DOWN | 0.00058970 | 0.045644 | 10.43 | 77.56 |
| 4 | atrazine CTD<br>00005450 | 0.00062691 | 0.045644 | 8.05 | 59.35 |
| 5 | medrysone HL60<br>DOWN | 0.00080205 | 0.045644 | 12.72 | 90.67 |

### Downregulation of DLAT Inhibits LC Cell Migration and Proliferation

In the previous analysis, we found that DLAT has the best prognostic value for 1, 2 and 3 years. Thus, we evaluated the function of DALT. We verified the expression level of DLAT in TCGA and GTEx database. DLAT is highly expressed in tumor tissues than in normal (Fig. 11A). And highly expressed DLAT is associated with poor prognosis (Fig. 11B). We first evaluated the transfection efficiency of the cells by qRT-PCR and found that the relative expression level of DLAT was significantly lower after shRNA transfection (Figure 11C). To further confirm the role of DLAT in invasion, transwell assays were performed. Our results showed that the invasion and migration rates of huh-7 and huh-1 cells transfected with shRNA were significantly lower than that of the control-transfected cells (Figure 11D). We performed CCK8 assays to detect the effect of DLAT knockdown on cell proliferation. After DLAT silencing, cell proliferation significantly decreased compared to control cells (Figure 11E). Colony formation assay also indicated that DLAT silencing significantly suppressed the growth of cells (Figure 11F). These data suggest that DLAT knockdown repressed the proliferative and invasive abilities of huh-7 and huh-1 cells.

**Figure 11.**
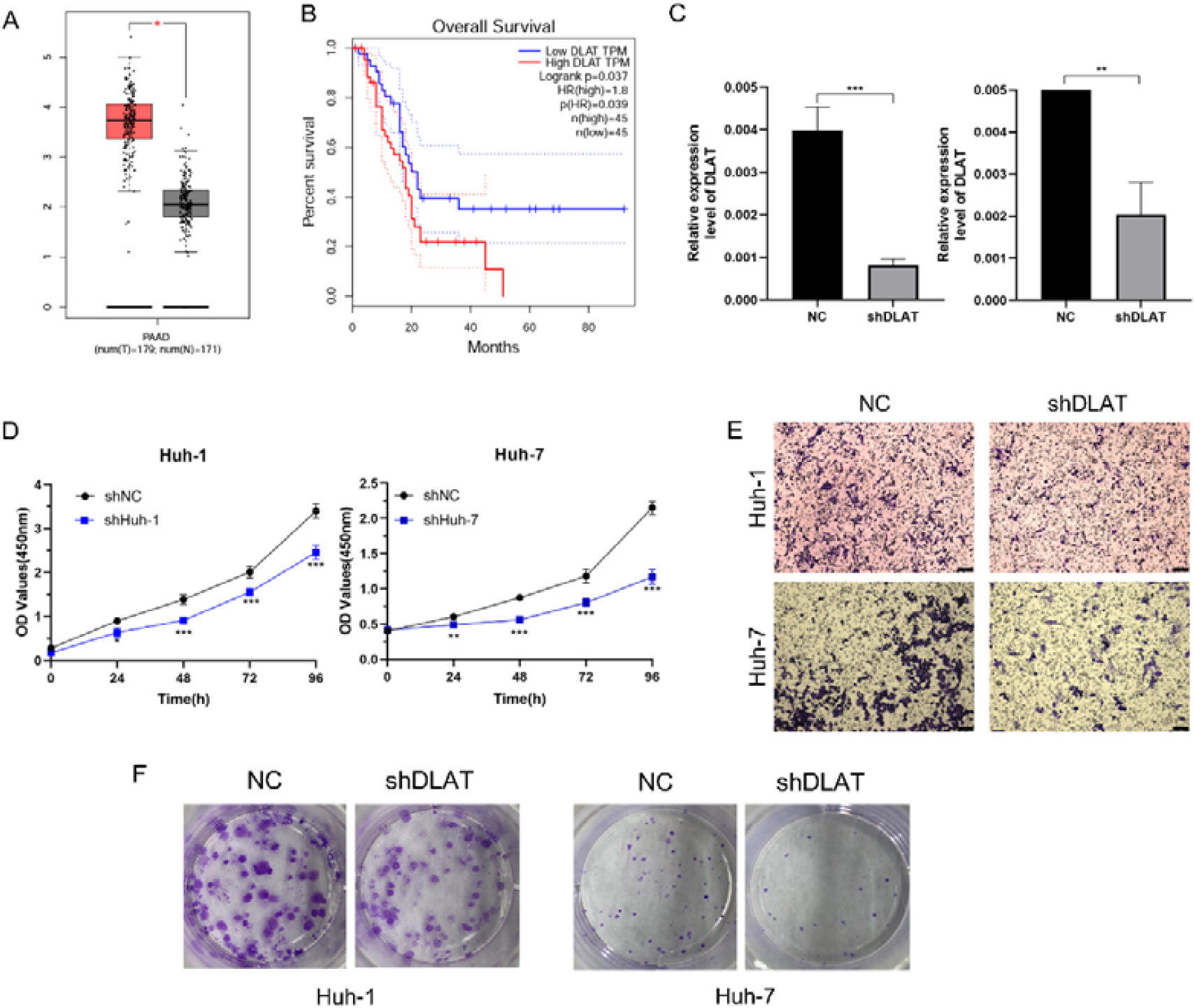
Functional experiments of DLAT **A** Expression level of DLAT in TCGA and GTEx database. **B** Overall survival of DLAT in TCGA. **C** Transfection efficiency was verified after transfection of DLAT or negative control shRNA. **D** LC cell viability was evaluated with CCK-8 assays at 0, 24, 48, 72, 96 h post-transfection. **E** Transwell assays were used to detect LC invasion and migration. Representative experiments are shown. **F** The number of LC cell colonies was reduced after DLAT knockdown.

### Potential therapeutic value of DLAT

We evaluated the correlation between the expression level of DLAT and half maximal inhibitory concentration (IC50) of the drugs. Among them, 9 pairs exhibited that drug resistance was associated with the expression level of DLAT, including GSK-2126458 (Cor = −0.43, p < 0.001), apitolisib (Cor = −0.387, p = 0.002), staurosporine (Cor = −0.38, p = 0.003), AT-13387 (Cor = 0.369, p < 0.004), everolimus (Cor = −0.364, p = 0.004), teglarinad (Cor = 0.362, p = 0.005), CUDC-305 (Cor = 0.34, p = 0.008), belinostat (Cor = 0.338, p = 0.008) and daporinad (Cor = 0.336, p = 0.009) (Figure 12).

**Figure 12.**
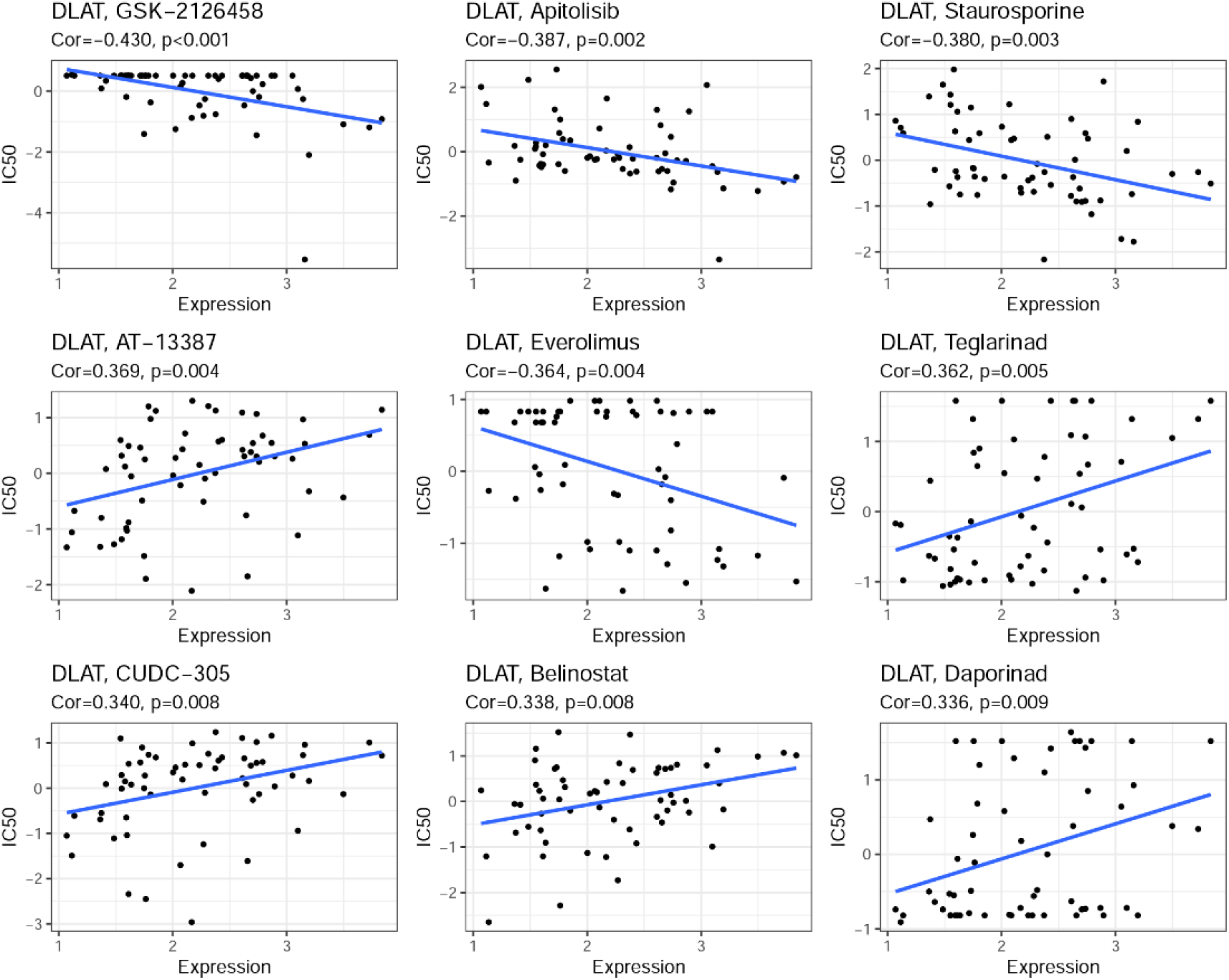
Correlation between the expression level of DLAT and half maximal inhibitory concentration (IC50) of the drugs.

## Discussion

The mRNA levels of 13 currently known cuproptosis-related genes in LC and normal tissues were compared in this study, and the majority of them were shown to be differently expressed. We used Cox univariate analysis and LASSO Cox regression analysis to create a 3-gene risk signature to further investigate the prognostic relevance of these cuproptosis-related regulators. The DEGs between the low- and high-risk groups were linked to cancer-related pathways, according to functional studies. When we analyzed the immune cell infiltration and activated pathways in the low- and high-risk groups, we discovered that the high-risk group had fewer amounts of mast cells, NK cells, and cytolytic activity than the low-risk group Through various subroutines, many heavy metals can cause controlled cell death. Intracellular copper buildup causes mitochondrial lipoylated proteins to aggregate and Fe–S cluster proteins to destabilize, leading to a unique type of cell death known as cuproptosis, according to a recent study published in *science*[4]. Their findings not only support the idea that mitochondria are complex regulators of cell death, including copper-induced cell death, but they also call into question the widely held belief that oxidative stress is a key molecular mechanism of metal toxicity [9–11]. How cuproptosis-related genes interact in LC and if they are linked to patient survival time are unknown. Our research discovered that a signature containing three cuproptosis-related genes (DLAT, LIPT1, and ATP7A) might predict OS in LC patients. The mitochondrial tricarboxylic acid (TCA) cycle is linked to dihydrolipoamide S-acetyltransferase (DLAT), which causes proteotoxic stress and cuproptosis[4]. DLAT is linked to cancer progression, chemoresistance, and cancer therapy in a variety of malignancies, including liver cancer, gastric cancer, prostate cancer, lymphocytic leukemia, and others[12–15]. We discovered it increased in liver cancer tissues as a cuproptosis-related gene. Furthermore, the DLAT risk model gave us some insight into possible prognostic and therapeutic targets for liver cancer. Lipoic acid is transferred from the H-protein of the glycine cleavage pathway to the E2 subunits of 2KDHs by lipoyltransferase-1 (LIPT1). LIPT1 mutations also cause mitochondrial disorders by disrupting lipoic acid metabolism[16]. LIPT1 is a lipoic acid pathway-related enzyme that is linked to mitochondrial respiration, implying a close relationship with the TCA cycle, which results in copper-induced cell death. ATP7A is a Cu-ATPase that transports copper from enterocytes into the blood via a transporter that travels to the basolateral membrane[17, 18]. The mutation of ATP7A causes a buildup of copper in enterocytes. Copper shortage is caused by this impairment in several connective tissues[19]. Menke’s disease-associated mice revealed increased intracellular copper accumulation in Tsvetkov’s study. In vivo, this copper buildup causes cell death[4]. In conclusion, three genes in the prognostic model (DLAT, LIPT1, and ATP7A) were found to be cuproptosis promoters. The risk model created by the three genes can predict prognosis. However, more research on how these genes interact with one another during cuproptosis is needed.

Cuproptosis is a newly discovered type of normal cell death that will require additional research in the future. Tsvetkov et al., for example, found that FDX1 deletion reduced DLAT lipoylation, implying that copper-induced cell death is dependent on protein lipoylation. They did not, however, explain the involvement of FDX1 in the lipoylation process. Similarly, the authors showed that copper binding to lipoylated DLAT increased disulfide-dependent DLAT oligomerization, but they did not explain the connection between copper and disulfide bond formation. Despite this, the authors provided a fresh perspective on the critical relationship between copper-induced mortality and cell mitochondrial metabolism. We compared DEGs between risk groups and discovered that DEGs were mostly engaged in channel activity. It’s in accordance with the cuproptosis personality. Many cancer-related pathways were elevated in KEGG pathway analysis, including the PI3K-AKT signaling system, the wnt signaling route, and the hippo signaling pathway. As a result, it’s fair to believe that cuproptosis is involved in cancer development.

In a process known as cancer immunosurveillance, the immune system may detect and eliminate emerging tumor cells, which serves as a crucial cancer defense. The immune system, on the other hand, can aid tumor growth. Cancer immunoediting refers to the immune system’s dual host-protective and tumor-promoting functions. Mast cells and NK cells were found to be relatively low in the high-risk group, whereas aDCs, iDCs, and Treg cells were abundant. This shows that the high-risk group’s immune function is impaired. The high levels of APC co-stimulation and MHC class I in the high-risk group suggest that the tumor microenvironment has been activated. However, in the high-risk group, cytolytic activity was low, indicating that T cell function was hindered, which is beneficial for tumor development[20].

Cuproptosis, particularly its mechanism in LC, is the subject of little current research. Our research found 12 genes that are differentially expressed in LC and their potential to regulate cuproptosis. We investigated the predictive usefulness of these cuproptosis-related genes in the preliminary stages and offered theoretical evidence for further research. We were unable to confirm if these regulators (which have been described in other studies) also play similar roles in cuproptosis pathways in LC due to a lack of data, and this question warrants additional investigation.

## Conclusion

In conclusion, our findings showed that cuproptosis is directly linked to LC since most cuproptosis-related genes were expressed differently in normal and LC tissues. Furthermore, in the TCGA cohorts, the score produced from our risk signature based on three cuproptosis-related genes was an independent risk factor for predicting OS. The differences in DEGs between the low-risk and high-risk groups were linked to tumor immunity. Our research identifies a new gene signature for predicting the prognosis of LC patients and lays the groundwork for future research into the link between cuproptosis-related genes and immunology in LC.

## Materials and methods

### Datasets

We used the TCGA database to get RNA sequencing (RNA-seq) data and clinical characteristics for LC patients up to May 13, 2022 (https://portal.gdc.cancer.gov/repository). The scale technique in the “limma” R package was used to standardize the gene expression profiles. Since these data were all available online with usage allowance, an extra ethical approval was not necessary.

### Identification of differentially expressed cuproptosis-related genes

From a previous article[4], we retrieved 13 cuproptosis-related genes, which are listed in Additional file 1. Before comparison, the expression data were standardized to fragment per kilobase million (FPKM) values. DEGs with a P value less than 0.05 were identified using the “limma” program. The DEGs are denoted as * if P < 0.05, ** if P < 0.01, and *** if P < 0.001. The Search Tool for the Retrieval of Interacting Genes (STRING), version 11.0 (https://string-db.org/), was used to create a PPI network for the DEGs.

### Development and validation of the cuproptosis-related gene prognostic model

We used Cox regression analysis to investigate the relationships between each gene and survival status in the TCGA cohort to determine the predictive efficacy of the cuproptosis-related genes. And the cut-off P-value was chosen at 0.01, and three survival-related genes (DLAT, LIPT1, ATP7A) were found for future investigation. To filter the potential genes and create the prognostic model, the LASSO Cox regression model (R package “glmnet”) was used. The three genes and their coefficients were eventually kept, and the penalty parameter (λ) was determined using the minimum criteria. The risk score formula was as follows: P = *∑*^7^_i_ *Xi* * *Yi* (*X*: coefficients, *Y*: gene expression level). According to the median risk score, the LC patients were separated into low- and high-risk subgroups, and the OS time was compared using Kaplan–Meier analysis. The “prcomp” function in the “stats” R package was used to compute PCA based on a 3-gene signature. A three-year ROC curve analysis was conducted using the R packages “survival”, “survminer”, and “time ROC”. And 70% of LC patients were randomly drawn to validate the model.

### Independent prognostic analysis of the risk score

The clinical information (age and grade) of patients in the TCGA cohort was retrieved. In our regression model, these variables were examined in conjunction with the risk score. The study used both univariate and multivariable Cox regression models.

### Functional enrichment analysis of the DEGs between the low- and high-risk groups

According to the median risk score, LC patients in the TCGA cohort were divided into two subgroups. DEGs were filtered across the low- and high-risk groups using particular criteria (|log_2_FC| ≥ 1 and FDR < 0.05). The “clusterProfiler” program was used to run GO and KEGG analyses based on these DEGs. The ssGSEA was used to determine the scores of invading immune cells and analyze the activity of immune-related pathways using the “gsva” program.

### Nomogram establishment based on risk score and clinical variables

We researched the relationship between DEG-based signature and clinical variables. In addition, combined with other clinical variables, we performed univariate and multivariate Cox regression analyses for exploring whether risk scores had an independent prognostic value in LC patients. Clinical variables and the DEG-based signature risk score were applied to establish a nomogram associated with outcome for evaluating the probability of 1-, 3-, and 5-year OS for LC patients. The concordance index (C-index) and calibration curve were performed to assess the predictive utility of the nomogram.

### Immune cell infiltration analysis

Mounting research confirmed that tumor cells immune infiltration was involved in cancer progression and correlated with prognosis. Therefore, we evaluated the infiltration level of immune cells between high-risk groups and low-risk groups by using CIBERSORT, CIBERSORT-ABS, QUANTISEQ, MCP-counter, XCELL, TIMER, and EPIC algorithms.

In order to predict the effect of immune checkpoint blockade therapy, we also explored the expression of several immune checkpoints. In addition, the TIMER database (https://cistrome.shinyapps.io/timer/) was used to identify the relationship between immune cells and DEGs and improve our understanding of the role of DEGs in LC.

### Drug sensitivity analysis

The correlation between DLAT and corresponding antitumor drug sensitivity in cancer cell lines were available from Genomics of Drug Sensitivity in Cancer (http://www.cancerrxgene.org/downloads). In addition, IC50, namely the concentration of an antitumor drug that is required to inhibit 50% cancer cells, was used to represent drug sensitivity.

### Clinical specimens

Between 2021 Jan. and 2022 May., liver cancer tissues and normal tissues were acquired from patients who were diagnosed with liver cancer and underwent surgery in the second affiliated hospital of Nanjing Medical University. Before using clinical materials, all patients signed an informed consent form. The ethics committee at Nanjing Medical University has approved the use of tissues in this study.

### Cell culture

The Chinese Academy of Sciences’ Type Culture Collection provided the liver cancer cell lines (Shanghai, China). All cells were grown at 37 °C in a humidified incubator in DMEM media with 10% fetal bovine serum (FBS; Gibco, Australia) and 1% penicillin– streptomycin with 5% CO_2_.

### RNA extraction and quantitative real-time PCR (qRT-PCR)

Trizol reagent (Invitrogen, USA) was used to isolate total RNA from tissues and cells according to the manufacturer’s procedure. HiScript II (Vazyme, China) was used to make cDNA, and qRT-PCR for mRNA and miRNA was done on an Applied Biosystems StepOne Plus Real-Time PCR machine or a LightCycler 480 (Roche, USA). The internal standard control for mRNA detection was GAPDH. The data was examined by comparing Ct values after each sample was reproduced three times. GeneCopoeia (Guangzhou, China) provided all PCR primers and shRNA of DLAT, which are presented in Additional file 2.

### Immunohistochemistry (IHC)

Protein expression was assessed by IHC on paraffin-embedded tissues. Xylene and 100 percent ethanol were used to treat tissue slides, followed by lower ethanol concentrations. Slides were blocked and stained with primary antibody after antigen retrieval, then secondary antibody incubation using the standard avidin biotinylated peroxidase complex procedure. Counterstaining was done with hematoxylin, and images were taken with an upright microscope system (Nikon, JAPAN). Image J was used to measure the integrated optical density (IOD), and AOD (Average Optical Density, AOD = IOD/area) was calculated.

### Transwell Migration

The invasion assay was conducted using 8-μm pore inserts coated with 30 μg of Matrigel (BD Biosciences). Cells were added to the coated filters. Additionally, cells were cultured in 6-well-plates and scraped with a 200-μl pipette tip. The cells were cultured in DMEM without FBS. Cell migration was photographed using an inverted microscope (OLYMPUS IX73, Olympus, Tokyo, Japan) at 0 and 24 h after injury.

### Cell Viability and Colony Formation Assays

Briefly, cells were seeded in 96-well-plates. After different incubation times, cell viability was measured with the Cell Counting Kit-8 (CCK-8, Dojindo, Kumamoto, Japan). Regarding colony formation experiment, 1,000 cells were seeded in cell culture plates and allowed to grow until visible colonies formed. Cell colonies were fixed with methanol, stained with crystal violet, and counted.

### Statistical analysis

The gene expression levels in normal liver and LC tissues were compared using single-factor analysis of variance, while categorical variables were compared using the Pearson chi-square test. We used the Kaplan–Meier method with a two-sided log-rank test to compare patient OS between subgroups. We employed univariate and multivariate Cox regression models to analyze the risk model’s independent prognostic efficacy. The Mann–Whitney test was performed to compare immune cell infiltration and immune pathway activation between the two groups. R software was used to conduct all statistical analyses (v4.0.2).

## Data Availability

All data produced in the present study are available upon reasonable request to the authors.
All data produced in the present work are contained in the manuscript.
All data produced are available online at TCGA database.

https://portal.gdc.cancer.gov/

## Abbreviations

LC: Liver cancer
DEG: Differentially expressed genes
TCGA: The Cancer Genome Atlas
LASSO: Least absolute shrinkage and selection operator
CCK8: Cell counting kit
OS: Overall survival
SEER: Surveillance Epidemiology End Results
LIPT1: Lipolytransferase 1
LIAS: Lipoyl synthase
DLD: Dihydrolipoamide dehydrogenase
PDH: Pyruvate dehydrogenase
ROS: Reactive oxygen species
RNA-seq: RNA sequencing
IC50: Half maximal inhibitory concentration
IHC: Immunohistochemistry
IOD: Integrated optical density
AOD: Average optical density
ROC: Receiver operating characteristic curve
AUC: Area under the ROC curve
GO: Gene ontology
KEGG: Kyoto Encyclopaedia of Genes and Genomes
TCA: Tricarboxylic acid

## Acknowledgements

None.

## Author contributions

X.D., Y.F. and C.Y. collected patient tissues, was one of the operators creating models, conceptualized the study, developed the technical tools necessary for data analysis, processed the data, wrote the initial manuscript and edited the manuscript. and they contribute equally to the article. Z.H. and S.H.J collected patient data, conceptualized the study and edited the manuscript. Y.X.J collected patient’s tissue samples. S.T. was one of the operators creating models, conceptualized the study and edited the manuscript. Z.Y.K. was one of the operators creating models, conceptualized the study and edited the manuscript. Z.G.W. conceptualized the study and was responsible for subject recruitment. C.D.X. conceptualized the study and edited the manuscript. L.K. and X.C.B. conceptualized the study, edited the manuscript and supervised the project.

## Funding

This study was supported in part by grants from the Health and Family Planning Commission of Jiangsu Province [#Z2020069], the Health and Family Planning Commission of Nanjing, Jiangsu Province[#YKK20172], and the Inheriting Studio of National Famous Old Chinese Medicine Experts-Zhu Yongkang(National Traditional Chinese Medicine Science and Education 2022 No. 75).

## Availability of data and materials

The datasets generated and analyzed during the current study are available from the corresponding author on reasonable request.

## Declarations

### Ethics approval and consent to participate

The studies involving human participants were reviewed and approved by the Ethics Committee of Nanjing Medical University. The patients/participants provided their written informed consent to participate in this study. Written informed consent was obtained from the individuals for the publication of any potentially identifiable images or data included in this article.

### Consent for publication

All authors have given consent for publication.

### Competing interests

The authors have no conflicts of interest to declare.

---

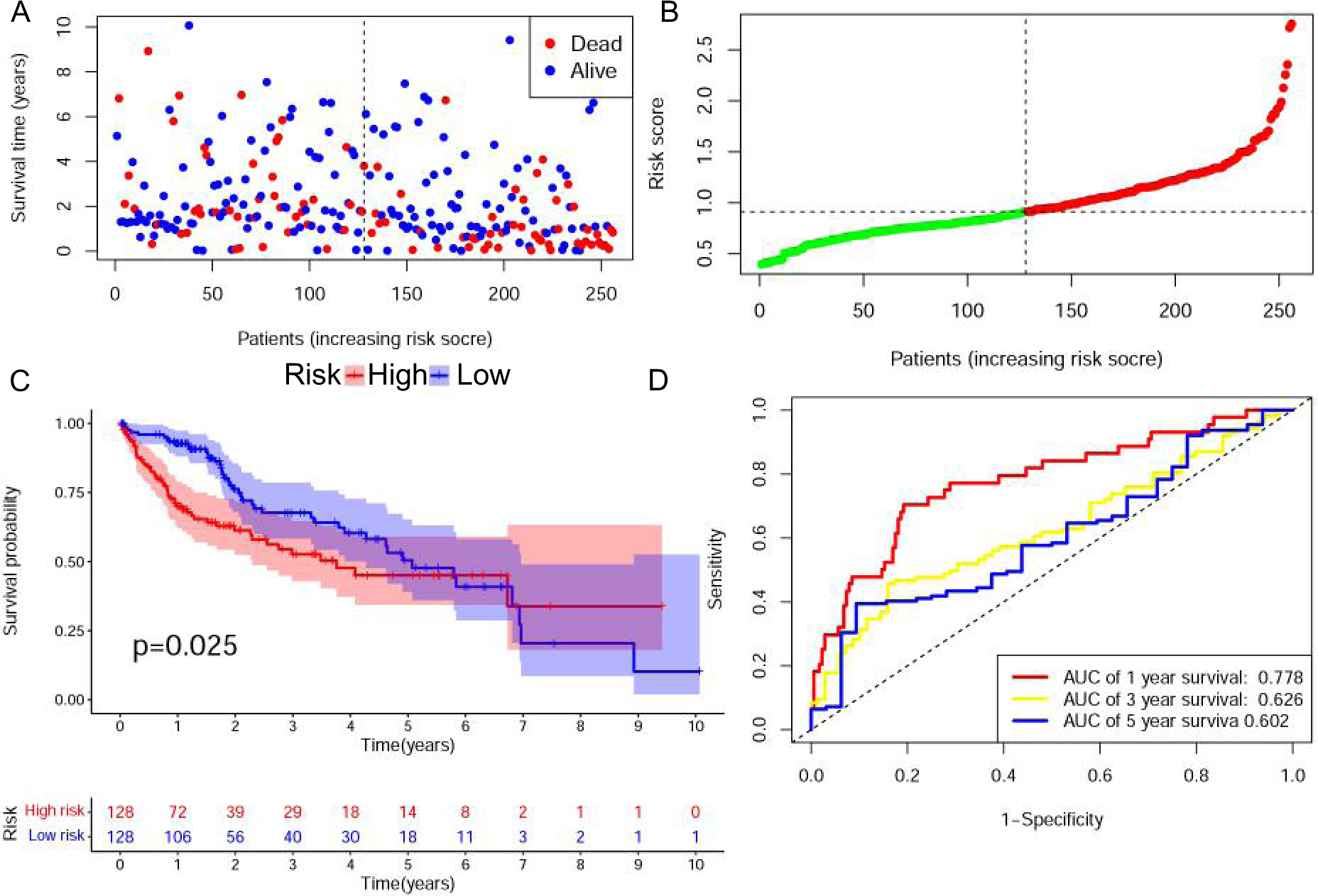

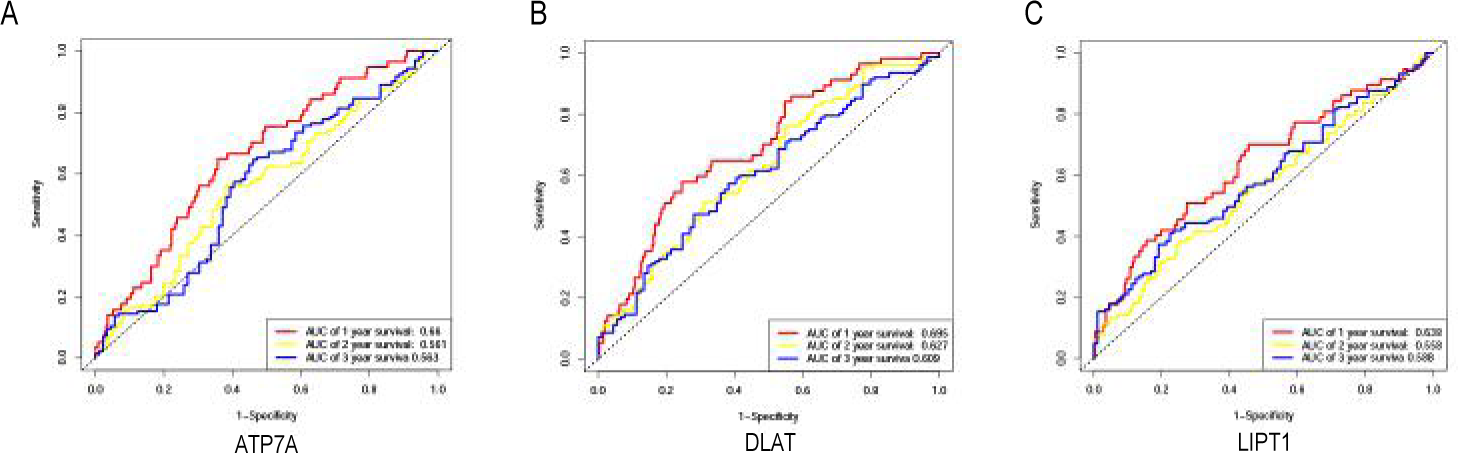

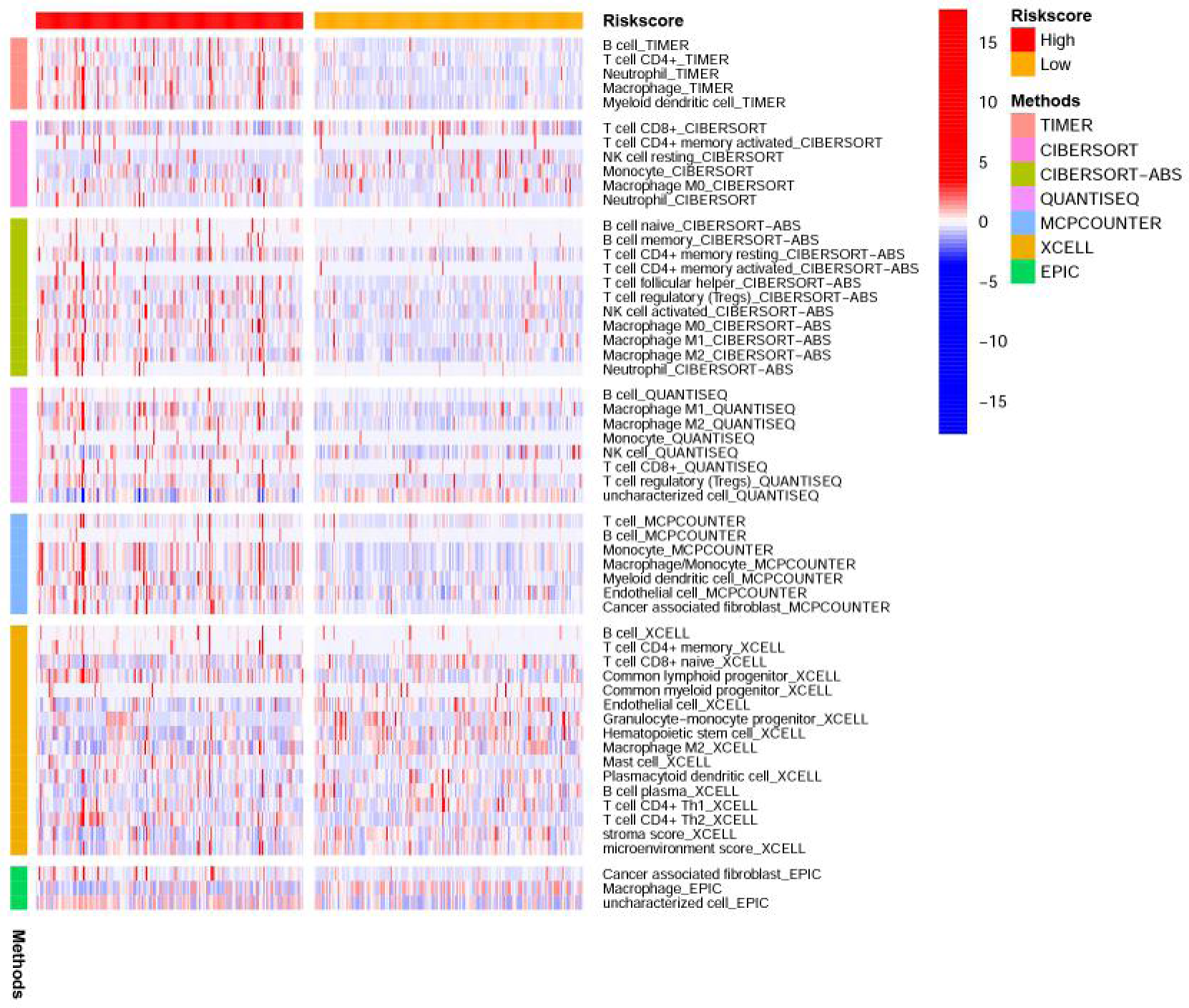

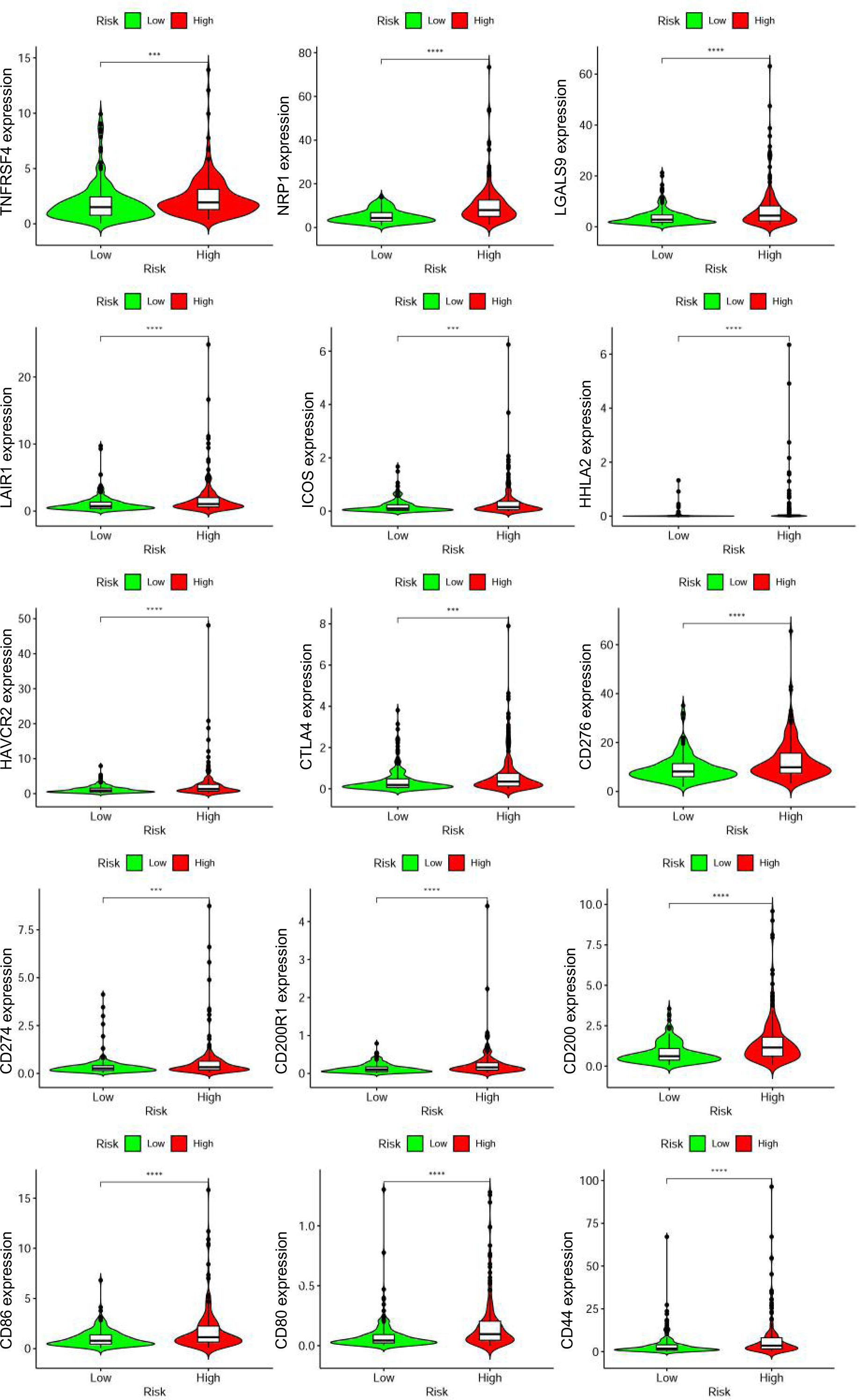

## Notes

### Competing Interest Statement

The authors have declared no competing interest.

### Author Declarations

The study used (or will use) ONLY openly available human data that were originally located at TCGA database.

